# Potential Role of Personal Protective Equipment Use in the Protection Against COVID-19 Infection Among Health Care Workers

**DOI:** 10.1101/2020.04.24.20070169

**Authors:** Wei Wang, Yuan-Zeng Min, Chun-Mei Yang, Hai-Ou Hong, Tian Xue, Yong Gao, Tengchuan Jin, Zhao-Hui Lu, Liang-Ming Zhang, Xueying Zheng, Sihui Luo, Wei Bao, Jian-Ping Weng

**Author notes:** **Corresponding Authors:** Jian-Ping Weng, MD, PhD, The First Affiliated Hospital of USTC, Division of Life Sciences and Medicine, University of Science and Technology of China, 17 Lujiang Road, Hefei, Anhui 230001, China and Wei Bao, MD, PhD, Department of Epidemiology, College of Public Health, University of Iowa, 145 North Riverside Drive, Room S431 CPHB, Iowa City, IA 52242, USA. These authors contributed equally to this work (Wei Wang, Yuan-Zeng Min, Chun-Mei Yang).

## Abstract

The coronavirus disease 2019 (COVID-19) pandemic has posed a major challenge for protecting health care workers (HCWs) against the infection. Use of personal protective equipment (PPE) in health care workplace is recommended as a high priority. In order to investigate the relationship between PPE use and the number of COVID-19 cases among HCWs, we conducted a molecular epidemiological study among 142 HCWs who were dispatched from Hefei to work in Wuhan and 284 HCWs who remained in Hefei, China; both provided care for patients with COVID-19. Nucleic acid testing and SARS-CoV-2 specific antibody (IgM, IgG, IgA) detection were performed to confirm SARS-CoV-2 infection among those HCWs. We also extracted publicly released data on daily number of COVID-19 cases among HCWs, daily number of HCWs who were dispatched to Hubei province since January 24, and daily production of PPE in China and daily demand and supply of PPE in Hubei province. Our laboratory testing confirmed that none of the 142 HCWs who were dispatched to work in Wuhan and 284 HCWs who remained in Hefei were infected by SARS-CoV-2. Consistent with these findings, as of April 15, 2020, none of the 42,600 HCWs who were successively dispatched to Hubei province since January 24, 2020 was reported to have COVID-19. These HCWs were provided with adequate supply of PPE as committed by their original institutions or provinces. In contrast, during the early phase of COVID-19 epidemic in Hubei province, a substantial shortage of PPE and an increasing number of COVID-19 infection among HCWs were reported. With the continuing increase in domestic production of PPE in China, the PPE supply started to meet and then exceed the demand. This coincided with a subsequent reduction in the number of reported COVID-19 cases among HCWs. In conclusion, our findings indicate that COVID-19 infection among HCWs could be completely prevented. Appropriate and adequate PPE might play a crucial role in protecting HCWs against COVID-19 infection.

## INTRODUCTION

Coronavirus disease 2019 (COVID-19), first reported in Wuhan, Hubei province, China in December 2019,^1-3^ has emerged as a pandemic worldwide. It is caused by a novel coronavirus,^4-6^ now knowns as the severe acute respiratory syndrome coronavirus 2 (SARS-CoV-2).^7^ As of April 18, 2020, the pandemic has affected more than 200 countries and regions around the world, with a total number of confirmed cases more than 2.1 million, of whom more than 139,515 people died due to COVID-19.^8^ The United States is now the country with the largest number of reported cases, comprising about one-fifth of all reported infections.^9^ Moreover, the global number of infected cases is rapidly increasing, posing a serious threat to global public health.^10^

As the SARS-CoV-2 spreads globally, a major challenge is ensuring successful protection of health care workers (HCWs) against COVID-19. HCWs are at high risk of COVID-19 infection because of their close contact with infected patients in a relatively closed environment. A recent report from Wuhan, China showed that 1496 local HCWs in Wuhan had confirmed COVID-19 from December 8, 2019 to March 8, 2020, with daily rate of cases in local HCWs 3 times higher than that in the general population.^11^ Thousands of COVID-19 cases or even deaths from COVID-19 have been reported among HCWs in other emerging epicenters, such as Italy,^12^ Spain, and the United States.

To protect HCWs against COVID-19 infection, use of personal protective equipment (PPE), such as masks, protective clothing, gloves, and goggles, in health care workplace has been recommended as a high priority.^13^ PPE is not only important for HCWs who directly provide care for patients with known COVID-19, but also for HCWs in general practice because there are undiagnosed patients or even asymptotic patients.^14^ However, limited availability or serious shortage of PPE, especially N95 masks and protective clothing, is reported worldwide.^15^ It is imperative to understand whether and to what extent PPE use protects HCWs against COVID-19.

So far, COVID-19 outbreak in Wuhan and throughout China has been temporarily controlled.^11^ During the early phase of COVID-19 outbreak, the rate of COVID-19 infection among local HCWs in Wuhan rapidly increased from December 8, 2019 to January 22, 2020 and peaked through February 1, 2020,^11^ which was accompanied by insufficient PPE supply and limited knowledge of this novel disease. In response, a series of multifaceted efforts has been implemented,^11^ including increasing PPE production and allocating nationwide PPE supply to Hubei province. Moreover, from January 24 to March 8, 42,600 HCWs from 30 other provinces were successively dispatched to Hubei province, ensuring the adequacy of medical personnel in the epidemic area.^12^ Because most of these HCWs were not specialized infectious disease or respiratory disease, it was very challenging to ensure protecting them against COVID-19 infection. Most of these HCWs remained in Hubei province until the outbreak of COVID-19 in Hubei province was controlled. During this period, PPE availability was largely improved, and comprehensive PPE was widely used among HCWs.

We conducted a molecular epidemiological study to confirm SARS-Cov-2 infection status among two groups of HCWs (one group were dispatched from Hefei to work in Wuhan and the other group remained in Hefei). We analyzed publicly released nationwide and regional data to depict the secular trends in PPE availability and the number of reported COVID-19 cases among HCWs. We hypothesize that PPE use may play an important role in the prevention of COVID-19 infection among HCWs.

## METHODS

The study protocol was approved by the Institutional Review Board at the University of Science and Technology of China (USTC).

### Data sources for the analyses of PPE availability and COVID-19 cases among HCWs in Hubei province

We extracted daily number of confirmed COVID-19 cases of HCWs during the early phase of COVID-19 response in China (as of February 14, 2020) from the data of epidemiological survey of 72,314 COVID-19 cases, which were reported by Chinese Center for Disease Control and Prevention.^16,17^ In addition, we obtained daily number of HCWs who were dispatched to work in Hubei province from January 24 to March 1, 2020, according to reports of National Health Commission of the People’s Republic of China (NHCPRC) and Health Commission of each of the 30 original provinces where the HCWs were dispatched from (**Supplementary Table 1**). We also obtained data about daily capacity of production of PPE (e.g., protective clothing) in China and daily demand and supply in Hubei province from websites of NHCPRC and the Health Commission of Hubei Provincial (HCHP). Data sources were listed in **Supplementary Table 2-4**.

### Molecular epidemiological study

#### Study population

In this molecular epidemiological study, we included two groups of HCWs, including HCWs who were dispatched from Hefei to work in Wuhan and those who remained in Hefei. Hefei is the capital city of Anhui province, which is around 235 miles away from Wuhan. On February 13, 2020, 142 HCWs (49 males and 93 females) from Hefei were dispatched to Wuhan, then took over the designated Sixth Special Ward for patients with severe conditions and worked there for 35 days. HCWs were called from multiple clinical departments, including ICU, infectious disease department, and other departments, because of the need of multidisciplinary care for COVID-19 patients with multiple organ infections and/or comorbid chronic diseases, such as hypertension, coronary heart disease, diabetes and malignant tumors. When treating COVID-19 patients, these HCWs were equipped with appropriate and adequate PPE including protective clothing, masks, gloves, goggles and face shields. After the COVID-19 in Wuhan was controlled, they returned to Hefei on March 18, 2020 and were quarantined for 14 days. Throat swab samples and blood samples were collected on March 25, 2020, which was during the quarantine period. All HCWs in this group were released from quarantine on April 1, 2020. For comparison, we also collected throat swab samples and blood samples among 284 local HCWs in Hefei who provided care for COVID-19 cases but were not dispatched to work in Wuhan.

#### Nucleic acid testing

Throat swab samples were subjected to Real-Time Fluorescent RT-PCR to test for SARC-CoV-2 viral RNA. Briefly, the swabs were incubated in a water bath at 56 °C for 30 minutes. The total RNAs were then extracted with magnetic beads by automatic nucleic acid extractor (Jiangsu Shuoshi) and subsequently added into the amplification tubes. RT-PCR was performed using Real-Time Fluorescent RT-PCR Kit (BGI, sensitivity: 100 copies/mL) or The DaAn Gene nCov RNA Kit (DAAN GENE, sensitivity: 500 copies/mL) on a Slan 96P Real Time PCR System (Shanghai Hongshi Medical Technology Co., Ltd).

#### SARS-CoV-2 specific antibody detection

Serum SARS-CoV-2 specific antibody levels were measured with chemiluminescent kits (Kangrun Biotech) for IgA (Doc no. KR/CE-01-B10, Revision A/0), IgG (Doc no. KR/CE-02-B10, Revision A/0) and IgM (Doc no. KR/CE-03-B10, Revision A/0). Briefly, the NP or RBD viral antigens were coated to magnetic particles to catch SARS-CoV-2 specific IgA, IgM and IgG in patient sera. Then a second antibody that recognizes IgA, IgM or IgG and is conjugated with acridinium (which can react with substrates to generate a strong chemiluminescence) was added for detection of IgA, IgM and IgG, respectively. The detected chemiluminescent signal over background signal was calculated as relative light units (RLU), COI was the ratio of RLU to statistically determined cut-off (criterion). Serum samples were collected by centrifugation of whole blood in test tubes at room temperature for 15 min. Prior to testing, a denaturant solution was added to each serum to a final concentration of 1% TNBP, 1% Triton X-100. After adequate mixing by inverting, the samples were incubated at 30°C for 4 hours to completely denature any potential viruses. Such solvent/detergent (1%TNBP + 1%Triton X-100) treatment is recommended by WHO guidelines on viral inactivation and removal procedures intended to assure the viral safety of human blood plasma products.^18^ Virus deactivated serum samples were then diluted 40 times with dilution buffer and subjected to testing at room temperature. Then RLU was measured using a fully automatic chemical luminescent immunoanalyzer, Kaeser 1000 (Kangrun Biotech, Guangzhou, China).

SARS-CoV-2 RBD-specific IgA, IgM and IgG antibodies were purified from a serum pool of recovering patients (published elsewhere) to be used as standards. The concentrations were determined using Bradford method (using bovine serum albumin protein as a standard). These antibodies were used to make a standard curve for each antibody detection kits to quantify the absolute antibody amounts in serum.

### Statistical analysis

We presented categorical variables among HCWs who were dispatched from Hefei to work in Wuhan and those who remained in Hefei as count (%). We used SPSS (version 19.0) for all the analyses.

## RESULTS

### Molecular epidemiological study of COVID-19 among HCWs

We tested 142 HCWs (49 males and 93 females), including physicians and nurses from the First Affiliated Hospital of USTC, who were dispatched to Hubei to fight against the outbreak of COVID-19. Only about a quarter of these HCWs were from ICU, infectious disease, respiratory medicine, emergency, anesthesiology departments, and the majority was from other departments who had relatively less experience in dealing with infectious diseases. Surprisingly, among them, both the nucleic acid tests and serological antibody tests showed that none of these HCWs was infected with SARS-CoV-2. The test of serological antibodies included IgA, IgM and IgG, which ruled out the possibility of asymptomatic infection.

We also performed the nucleic acid tests and serological antibody tests on 284 HCWs who remained in Hefei and did not work in Wuhan. Similarly, both the nucleic acid and serological antibody tests showed no SARS-CoV-2 infection among them (**Table 1**).

**Table 1.** Characteristics of HCWs who were dispatched to work in Wuhan and HCWs who remained to work in Hefei.

| **Variables** | **HCWs who worked in**<br>**Wuhan** | **HCWs who remained in**<br>**Hefei** |
| --- | --- | --- |
| **No. of participants** | 142 | 284 |
| **Age** |  |  |
| 20-29 | 40 (28.17%) | 90 (31.69%) |
| 30-39 | 72 (50.71%) | 144 (50.70%) |
| 40-49 | 27 (19.01%) | 42 (14.79%) |
| 50-59 | 3 (2.11%) | 8 (2.82%) |
| ≥60 | 0 (0.00%) | 0 (0.00%) |
| **Gender** |  |  |
| Female | 93 (65.49%) | 184 (64.79%) |
| Male | 49 (34.51%) | 100 (35.21%) |
| **Occupational type** |  |  |
| Nurse | 101 (71.13%) | 123 (43.31%) |
| Doctor | 38 (26.76%) | 87 (30.63%) |
| Others | 3 (2.11%) | 74 (26.06%) |
| **Original Department** |  |  |
| ICU | 15 (10.56%) | 4 (1.41%) |
| Infectious Diseases | 2 (1.41%) | 1 (0.35%) |
| Respiratory Medicine | 11 (7.75%) | 6 (2.11%) |
| Emergency | 8 (5.63%) | 12 (4.23%) |
| Anesthesiology | 3 (2.11%) | 4 (1.41%) |
| Others | 103 (72.54%) | 257 (90.49%) |

### Trends in PPE availability and COVID-19 cases among HCWs in Hubei province

A substantial shortage of PPE was reported during the early phase of COVID-19 epidemic in China. According to data reported by NHCPRC and the Ministry of Industry and Information Technology of the People’s Republic of China (MIITPRC), during the period from January 23 to February 1, the daily demand of PPE was on average 100,000 sets per day, while China’s domestic average capacity of production was less than 10,000 sets per day. During that period, PPE supply was heavily dependent on overseas imports and donations except domestic production. With the continuing increase in domestic production of PPE in China, the PPE supply started to meet and then exceed the demand. On February 7, 2020, Hubei province needed about 59,400 sets of PPE per day, while China’s daily production capacity reached 48,500 sets. Together with overseas imports and donations, the demand and supply were tightly balanced at that time. After March 6, 2020, daily production of PPE in China reached 500,000 sets, exceeding the daily demand (**Figure 1**).

**Figure 1.**
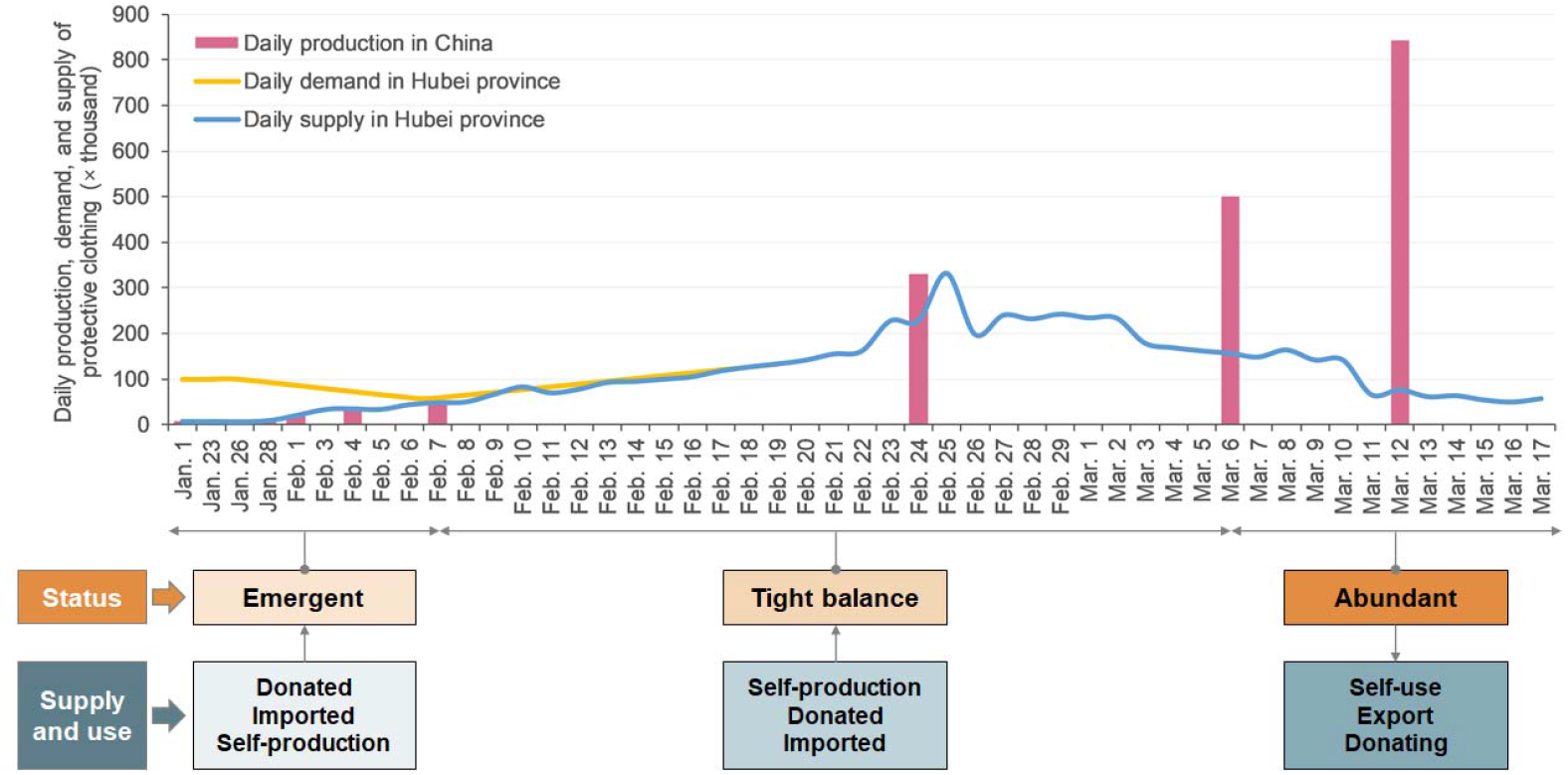
Secular trends in daily production of medical protective clothing in China and its daily demand and supply balance in Hubei province.

The number of confirmed COVID-19 infections among HCWs in China increased since the outbreak in December 2019, peaked on January 28, 2020, and then dropped gradually thereafter (**Figure 2**). As a strategy to provide sufficient HCWs to deal with the drastically increased number of COVID-19 patients and to compensate the reduced number of available HCWs in Hubei province due to COVID-19 infection and quarantine, 42,600 HCWs from other provinces were successively dispatched to work in major hospitals in Hubei province since January 24, 2020. As of April 16, 2020, none of the 42,600 HCWs was reported to have COVID-19 infection (**Figure 3**).

**Figure 2.**
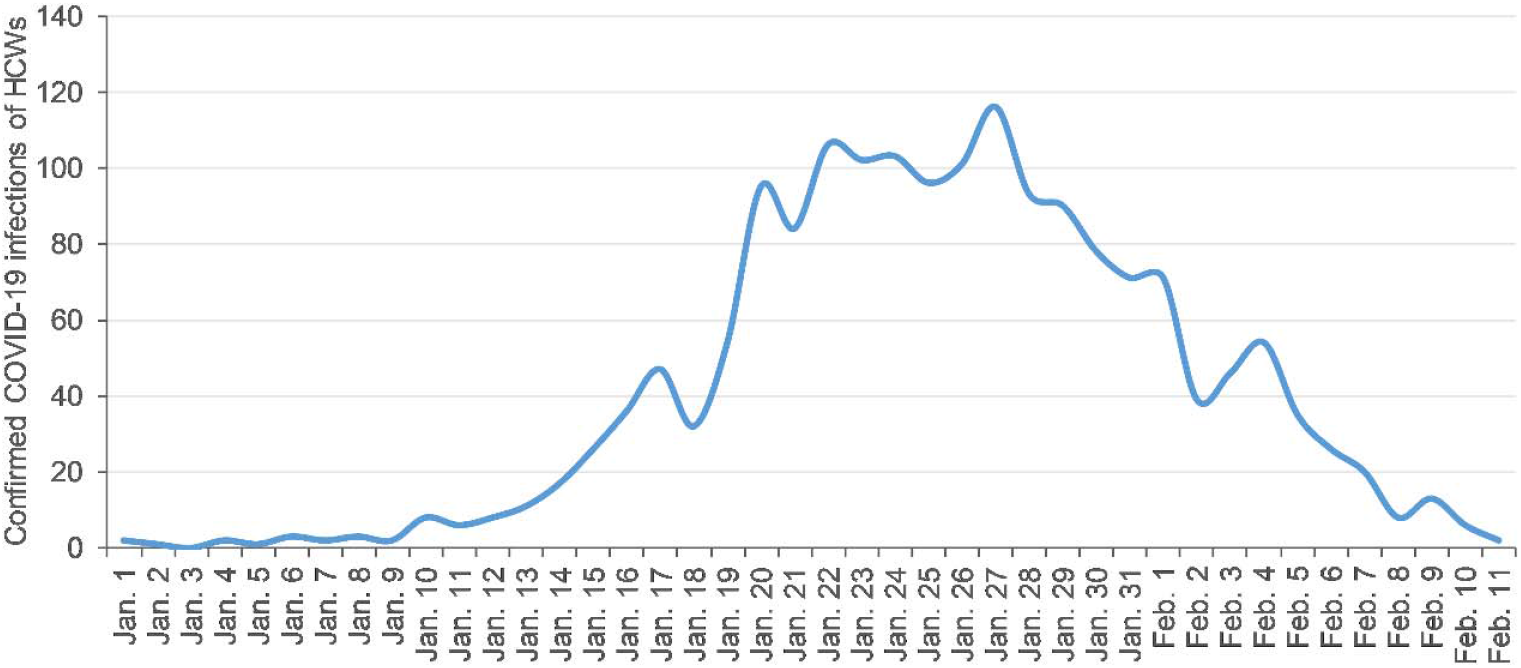
Secular trends in the number of confirmed COVID-19 cases among HCWs in China. Reproduced with permission from data in a previous report.^17^

**Figure 3.**
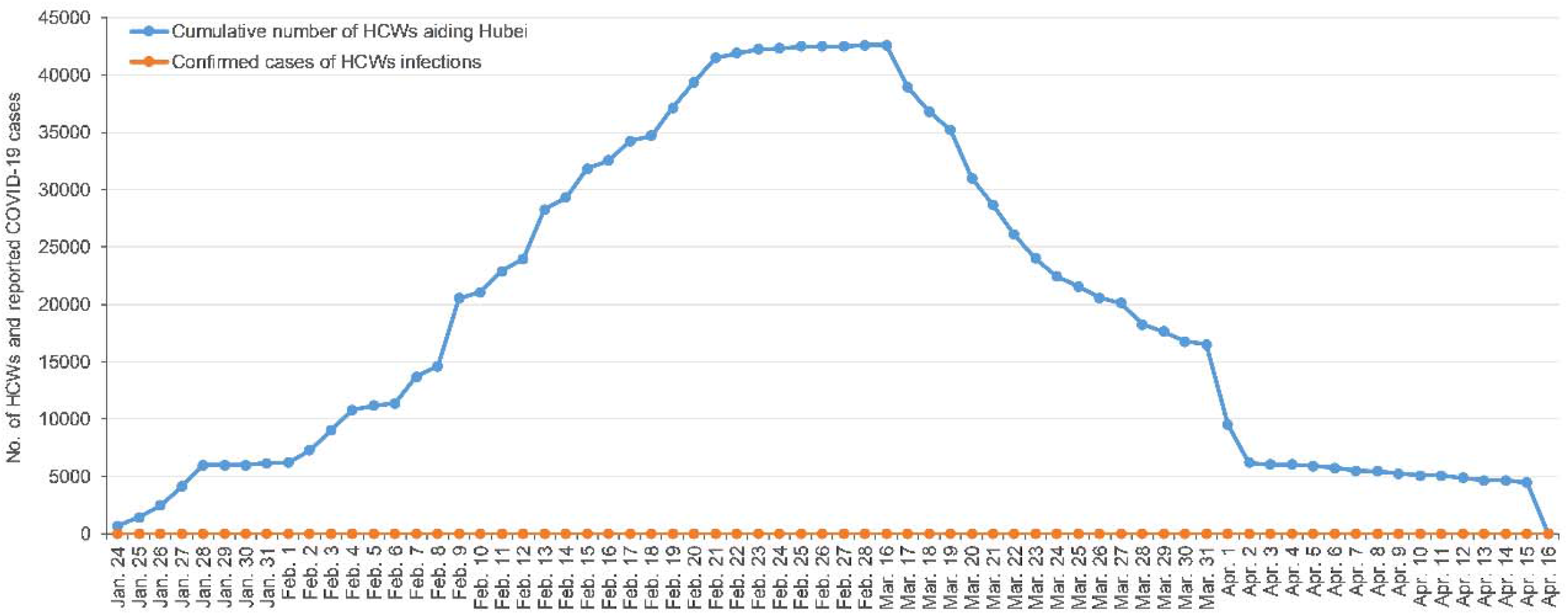
Secular trends in the number of HCWs who were dispatched to work in major hospitals in Hubei province and the number of cases of COVID-19 among those HCWs. A total of 42,600 HCWs from other provinces were successively dispatched to Hubei province from January 24 to March 1, 2020. After the epidemic of COVID-19 was controlled, they were successively withdrawn since March 17, 2020. No infection was reported among them until April 16, 2020.

## DISCUSSION

This molecular epidemiological study of two groups of HCWs confirmed that HCWs who were dispatched to work in Wuhan and those who remained in Hefei were not infected by SARS-CoV-2. The antibody testing including IgA, IgM and IgG was able to exclude any asymptomatic infections in the HCWs if any. Therefore, our findings supported the report about zero infection of COVID-19 among 42,600 HCWs who were successively dispatched to Hubei province to fight against COVID-19.^19^ Many HCWs worked in Hubei province for two months with high risk of contracting the virus. Along with more than 80,000 local HCWs in Hubei province, these HCWs had cured 55,987 COVID-19 patients in Hubei province. However, they remained free of COVID-19 infection.

Our analysis of national and regional PPE availability suggested a potential association between PPE use and reduction in COVID-19 infections among HCWs. During the early phase of the epidemic in Hubei province, data from both NHCPRC and MIITPRC showed an extreme shortage of the PPE and other medical supplies for HCWs in Hubei province. For example, the daily supply of protective clothing only met for less than 10% of the demand at that time. During that period, hundreds of HCWs were infected with COVID-19 in Hubei province, and the number of COVID-19 cases among HCWs in Hubei province accounted for over 90% of the cases among HCWs nationwide. Subsequently, with nationwide efforts to promote domestic production, the supply of PPE was gradually meeting the demand since early February 2020, which is in parallel with the decrease in the number of COVID-19 cases among HCWs. Since early March, the emergency state has been reversed to a surplus status. Coincidently, the rapid increase of COVID-19 infection among local HCWs in Wuhan from December 8, 2019 to January 22, 2020, peaked from January 23 to February 1, 2020, and then decreased from February 2 to March 8, 2020.^11^

An important measure to protect the HCWs to be dispatched to Hubei province was adequate supply of PPE, as committed and provided by their original institutions or provinces. This was crucial because of the limited supply of PPE in Hubei province at that time. For example, Jiangsu province, which dispatched the largest number of HCWs (n=2,802) to Hubei province, sent 545,800 pieces of medical supplies including 516,000 pieces of PPE, to Hubei province by February 22, 2020. Similar measures to ensure PPE supply were taken from other provinces which dispatched HCWs to Hubei province. The adequate supply and use of PPE were likely to play an important role in the successful protection of all HCWs dispatched to Hubei province with no COVID-19 infection among them.

It is noteworthy that not all the HCWs that were dispatched to Hubei province had expertise in infectious disease or respiratory disease. In the present study, the majority of HCWs that were dispatched from Hefei to work in Wuhan were from departments other than ICU, infectious diseases, or respiratory diseases. Although these HCWs were provided adequate PPE, they had limited experience dealing with infectious agents in their previous clinical work. If not protected, exposure to symptomatic or asymptomatic patients of COVID-19 could cause infections among some HCWs. However, even among them, no one was infected with SARS-CoV-2, which further strengthened the importance of adequate PPE supply and use for HCWs.

Our findings about a potential relation between PPE use and successful protection against COVID-19 among HCWs were in line with available, although still limited, evidence. SARS-CoV-2 is primarily transmitted through droplets and in some circumstances, through aerosols.^9,20^ Recently, a study found that surgical masks significantly reduced detection of coronavirus RNA in aerosols, with a trend toward reduced detection of coronavirus RNA in respiratory droplets, indicating that surgical masks could prevent transmission of human coronaviruses from symptomatic individuals.^21^ Nosocomial transmission of SARS-CoV-2 has been reported among hospitalized patients and HCWs.^22^ A recent study showed extensive environmental contamination by a patient with SARS-CoV-2 through respiratory droplets and fecal shedding, which suggests the environment as a potential medium of transmission.^23^ Therefore, in order to block the various spreading routes of the virus, application of comprehensive PPE including masks, protective clothing, gloves, goggles and face shield, etc. is warranted to provide effective protection for HCWs.

This study has important clinical implications. Although HCWs often accept increased risk of infection, as part of their chosen profession, any possible effort must be taken to minimize their risk of COVID-19 infection. The first wave of COVID-19 outbreak in Wuhan and throughout China has been successfully controlled, with the lockdown of Wuhan and a series of multifaceted interventions.^11^ However, emerging epicenters around the world, such as Italy,^24^ Spain, and the United States,^9^ are currently experiencing similar epidemiologic curve as what happened in January and early February in Wuhan, China. Therefore, learning experience from China may help inform policy making in other countries and regions. The laboratory-confirmed zero infection of COVID-19 in this study offers a promising message to the HCWs that their risk might be greatly reduced or even eliminated with appropriate and adequate PPE use.

This study has several limitations. First, like other observational studies, the present study could not establish causality between PPE use and the successful prevention of COVID-19 infection among HCWs. Due to ethical consideration, it is impossible to conduct experimental studies to compare infections between HCWs wearing and not wearing PPE. Only historical data of different periods could be used to test our hypothesis. Second, PPE consists of various items such as protective clothing, N95 masks, surgical masks, gloves, and goggles, etc. This study only presents the data of medical protective clothing because protective clothing is representative of PPE production and supply in China. Third, we were not able to test all the 42,600 HCWs who were dispatched to Hubei province. Finally, we did not have information about the HCWs’ personal prevention awareness and adherence at individual level which may also influence the infection among HCWs.

In conclusion, our laboratory-confirmed findings of zero infection indicate that COVID-19 infection among HCWs could be completely prevented. Appropriate and adequate PPE might play a crucial role in protecting HCWs against COVID-19 infection. Therefore, it is critical and urgent to ensure sufficient PPE in health care workplace to protect HCWs from COVID-19 infection while they are fighting against the pandemic.

## Data Availability

The data availability are included in our supplementary information.

## Contributors

WW, YZM, CMY, WB, and JPW designed the study. WW, YZM, and CMY collected the data. WW and YZM analysed the data. WW, YZM, CMY, WB, and JPW interpreted the data. WW wrote the first draft. WW, YZM, and CMY contributed equally to this work. All authors contributed to the final draft.

## Acknowledgments

We are grateful to the health care workers for their contributions to caring patients with COVID-19 and their participation in this study. This study was supported by the Fundamental Research Funds for the Central Universities (YD9110004001 and YD9110002002).

---

**Supplementary Table 1.**
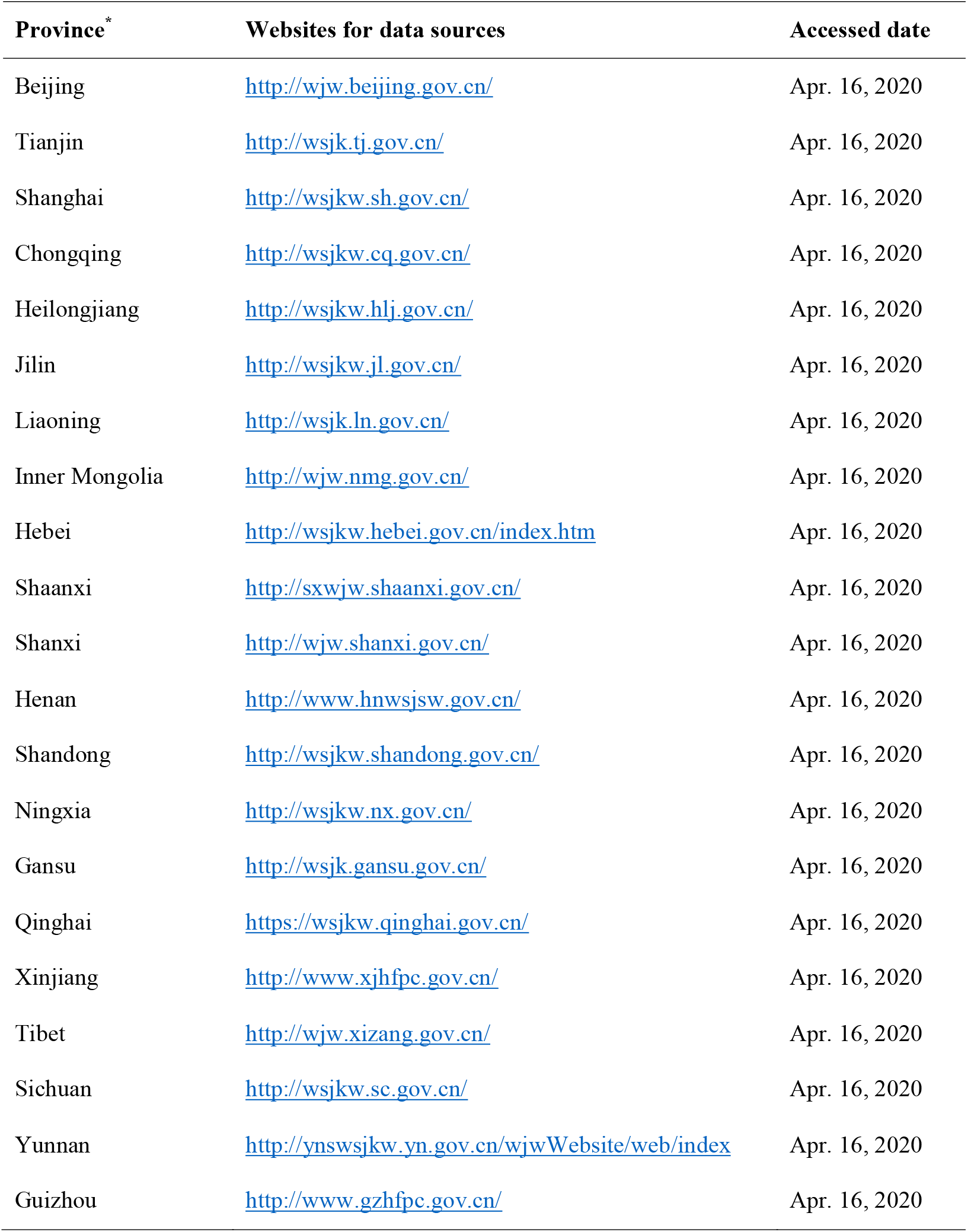
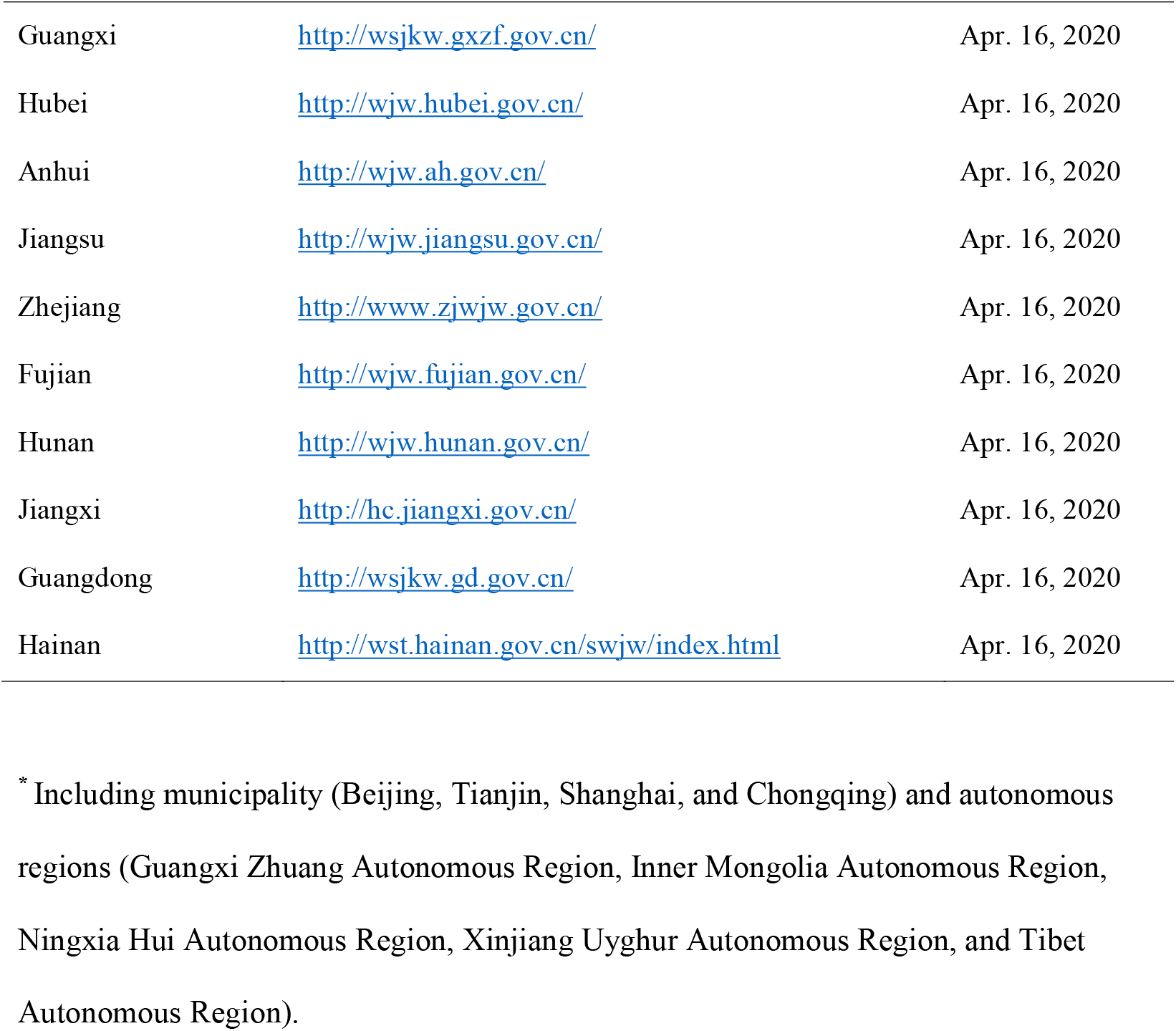
Data sources for the number of health care workers who were dispatched to Hubei province, their departure date, and zero infection report.

**Supplementary Table 2.** Data sources for daily production of medical protective clothing in China.

| **Date** | **Number (× thousand)** | **Websites for data sources** | **Release date** | **Accessed date** |
| --- | --- | --- | --- | --- |
| Before Jan. 23, 2020 | 7 | http://wjw.hubei.gov.cn/fbjd/dtyw/202002/t20200209_2022202.shtml | Feb. 9, 2020 | Apr. 14, 2020 |
| Jan. 28, 2020 | 8.7 | http://www.nhc.gov.cn/xcs/fkdt/202002/8f01fbf4982b4c4ca6ef690d3ebfee74.shtml | Feb. 5, 2020 | Apr. 14, 2020 |
| Feb. 1, 2020 | 20 | http://www.gov.cn/xinwen/2020-02/02/content_5473932.htm | Feb. 2, 2020 | Apr. 14, 2020 |
| Feb. 4, 2020 | 31.6 | http://www.nhc.gov.cn/xcs/fkdt/202002/8f01fbf4982b4c4ca6ef690d3ebfee74.shtml | Feb. 5, 2020 | Apr. 14, 2020 |
| Feb. 7, 2020 | 48.5 | http://wjw.hubei.gov.cn/fbjd/dtyw/202002/t20200209_2022202.shtml | Feb. 9, 2020 | Apr. 14, 2020 |
| Feb. 24, 2020 | 330 | http://www.scio.gov.cn/xwfbh/xwbfbh/wqfbh/42311/42592/index.htm | Feb. 25, 2020 | Apr. 14, 2020 |
| Mar. 6, 2020 | 500 | http://www.scio.gov.cn/xwfbh/xwbfbh/wqfbh/42311/42682/index.htm | Mar. 6, 2020 | Apr. 14, 2020 |
| Mar. 12, 2020 | 842 | http://www.nhc.gov.cn/xcs/fkdt/202003/9df571ff7f144bc69f63bbb636fe2b2d.shtml | Mar. 12, 2020 | Apr. 14, 2020 |

**Supplementary Table 3.** Data sources for the daily demand of medical protective clothing (MPC) in Hubei province.

| **Date** | **Number (× thousand)** | **Websites for data sources** | **Release date** | **Accessed date** |
| --- | --- | --- | --- | --- |
| As of Jan. 26, 2020 | 100 | http://www.scio.gov.cn/xwfbh/xwbfbh/wqfbh/42311/42478/index.htm | Jan. 26, 2020 | Apr. 14, 2020 |
| Feb. 6, 2020 | 59.9 | http://wjw.hubei.gov.cn/bmdt/ztzl/fkxxgzbdgrfyyq/xxfb/202002/t20200208_2021729.shtml | Feb. 8, 2020 | Apr. 14, 2020 |
| Feb. 7, 2020 | 59.4 | http://wjw.hubei.gov.cn/bmdt/ztzl/fkxxgzbdgrfyyq/xxfb/202002/t20200209_2022197.shtml | Feb. 9, 2020 | Apr. 14, 2020 |

**Supplementary Table 4.** Data sources for daily supply of medical protective clothing (MPC) in Hubei province.

| **Supply date** | **Number** | **Websites for data sources** | **Release date** | **Accessed date** |
| --- | --- | --- | --- | --- |
| Before Jan. 1, 2020 | 7000 | http://wjw.hubei.gov.cn/fbjd/dtyw/202002/t20200209_2022202.shtml | Feb. 9, 2020 | Apr. 14, 2020 |
| Jan. 28, 2020 | 8700 | http://www.nhc.gov.cn/xcs/fkdt/202002/8f01fbf4982b4c4ca6ef690d3ebfee74.shtml | Feb. 5, 2020 | Apr. 14, 2020 |
| Feb. 1, 2020 | 20000 | http://www.gov.cn/xinwen/2020-02/02/content_5473932.htm | Feb. 2, 2020 | Apr. 14, 2020 |
| Feb. 3, 2020 | 32965 | http://wjw.hubei.gov.cn/bmdt/ztzl/fkxxgzbdgrfyyq/xxfb/202002/t20200204_2018839.shtml | Feb. 4, 2020 | Apr. 14, 2020 |
| Feb. 4, 2020 | 34807 | http://wjw.hubei.gov.cn/bmdt/ztzl/fkxxgzbdgrfyyq/xxfb/202002/t20200205_2019348.shtml | Feb. 5, 2020 | Apr. 14, 2020 |
| Feb. 5, 2020 | 33640 | http://wjw.hubei.gov.cn/bmdt/ztzl/fkxxgzbdgrfyyq/xxfb/202002/t20200206_2019934.shtml | Feb. 6, 2020 | Apr. 14, 2020 |
| Feb. 6, 2020 | 43950 | http://wjw.hubei.gov.cn/bmdt/ztzl/fkxxgzbdgrfyyq/xxfb/202002/t20200207_2020654.shtml | Feb. 7, 2020 | Apr. 14, 2020 |
| Feb. 7, 2020 | 48520 | http://wjw.hubei.gov.cn/bmdt/ztzl/fkxxgzbdgrfyyq/xxfb/202002/t20200208_2021482.shtml | Feb. 8, 2020 | Apr. 14, 2020 |
| Feb. 8,<br>2020 | 49910 | http://wjw.hubei.gov.cn/bmdt/ztzl/fkxxgzbdgrfyyq/xxfb/202002/t20200209_2022007.shtml | Feb. 9,<br>2020 | Apr. 14,<br>2020 |
| Feb. 9,<br>2020 | 66429 | http://wjw.hubei.gov.cn/bmdt/ztzl/fkxxgzbdgrfyyq/xxfb/202002/t20200210_2022649.shtml | Feb. 10,<br>2020 | Apr. 14,<br>2020 |
| Feb. 10,<br>2020 | 83096 | http://wjw.hubei.gov.cn/bmdt/ztzl/fkxxgzbdgrfyyq/xxfb/202002/t20200211_2023577.shtml | Feb. 11,<br>2020 | Apr. 14,<br>2020 |
| Feb. 11,<br>2020 | 69992 | http://wjw.hubei.gov.cn/bmdt/ztzl/fkxxgzbdgrfyyq/xxfb/202002/t20200212_2024717.shtml | Feb. 12,<br>2020 | Apr. 14,<br>2020 |
| Feb. 12,<br>2020 | 77920 | http://wjw.hubei.gov.cn/bmdt/ztzl/fkxxgzbdgrfyyq/xxfb/202002/t20200213_2025625.shtml | Feb. 13,<br>2020 | Apr. 14,<br>2020 |
| Feb. 13,<br>2020 | 92720 | http://wjw.hubei.gov.cn/bmdt/ztzl/fkxxgzbdgrfyyq/xxfb/202002/t20200214_2027241.shtml | Feb. 14,<br>2020 | Apr. 14,<br>2020 |
| Feb. 14,<br>2020 | 95000 | http://wjw.hubei.gov.cn/bmdt/ztzl/fkxxgzbdgrfyyq/xxfb/202002/t20200215_2028437.shtml | Feb. 15,<br>2020 | Apr. 14,<br>2020 |
| Feb. 15,<br>2020 | 100000 | http://wjw.hubei.gov.cn/bmdt/ztzl/fkxxgzbdgrfyyq/xxfb/202002/t20200216_2038783.shtml | Feb. 16,<br>2020 | Apr. 14,<br>2020 |
| Feb. 16,<br>2020 | 105240 | http://wjw.hubei.gov.cn/bmdt/ztzl/fkxxgzbdgrfyyq/xxfb/202002/t20200217_2039477.shtml | Feb. 17,<br>2020 | Apr. 14,<br>2020 |
| Feb. 17,<br>2020 | 117870 | http://wjw.hubei.gov.cn/bmdt/ztzl/fkxxgzbdgrfyyq/xxfb/202002/t20200218_2095459.shtml | Feb. 18,<br>2020 | Apr. 14,<br>2020 |
| Feb. 18,<br>2020 | 126660 | http://wjw.hubei.gov.cn/bmdt/ztzl/fkxxgzbdgrfyyq/xxfb/202002/t20200219_2130720.shtml | Feb. 19,<br>2020 | Apr. 14,<br>2020 |
| Feb. 19,<br>2020 | 133470 | http://wjw.hubei.gov.cn/bmdt/ztzl/fkxxgzbdgrfyyq/xxfb/202002/t20200220_2142168.shtml | Feb. 20,<br>2020 | Apr. 14,<br>2020 |
| Feb. 20,<br>2020 | 141640 | http://wjw.hubei.gov.cn/bmdt/ztzl/fkxxgzbdgrfyyq/xxfb/202002/t20200221_2143831.shtml | Feb. 21,<br>2020 | Apr. 14,<br>2020 |
| Feb. 21<br>2020 | 155330 | http://wjw.hubei.gov.cn/bmdt/ztzl/fkxxgzbdgrfyyq/xxfb/202002/t20200222_2145012.shtml | Feb. 22,<br>2020 | Apr. 14,<br>2020 |
| Feb. 22,<br>2020 | 161940 | http://wjw.hubei.gov.cn/bmdt/ztzl/fkxxgzbdgrfyyq/xxfb/202002/t20200223_2145604.shtml | Feb. 23,<br>2020 | Apr. 14,<br>2020 |
| Feb. 23,<br>2020 | 227953 | http://wjw.hubei.gov.cn/bmdt/ztzl/fkxxgzbdgrfyyq/xxfb/202002/t20200224_2146206.shtml | Feb. 24,<br>2020 | Apr. 14,<br>2020 |
| Feb. 24,<br>2020 | 228490 | http://wjw.hubei.gov.cn/bmdt/ztzl/fkxxgzbdgrfyyq/xxfb/202002/t20200225_2146942.shtml | Feb. 25,<br>2020 | Apr. 14,<br>2020 |
| Feb. 25,<br>2020 | 332572 | http://wjw.hubei.gov.cn/bmdt/ztzl/fkxxgzbdgrfyyq/xxfb/202002/t20200226_2153201.shtml | Feb. 26,<br>2020 | Apr. 14,<br>2020 |
| Feb. 26,<br>2020 | 198250 | http://wjw.hubei.gov.cn/bmdt/ztzl/fkxxgzbdgrfyyq/xxfb/202002/t20200227_2160641.shtml | Feb. 27,<br>2020 | Apr. 14,<br>2020 |
| Feb. 27,<br>2020 | 240554 | http://wjw.hubei.gov.cn/bmdt/ztzl/fkxxgzbdgrfyyq/xxfb/202002/t20200228_2161418.shtml | Feb. 28,<br>2020 | Apr. 14,<br>2020 |
| Feb. 28,<br>2020 | 232540 | http://wjw.hubei.gov.cn/bmdt/ztzl/fkxxgzbdgrfyyq/xxfb/202002/t20200229_2162521.shtml | Feb. 29,<br>2020 | Apr. 14,<br>2020 |
| Feb. 29,<br>2020 | 242955 | http://wjw.hubei.gov.cn/bmdt/ztzl/fkxxgzbdgrfyyq/xxfb/202003/t20200301_2164983.shtml | Mar. 1,<br>2020 | Apr. 14,<br>2020 |
| Mar. 1,<br>2020 | 234950 | http://wjw.hubei.gov.cn/bmdt/ztzl/fkxxgzbdgrfyyq/xxfb/202003/t20200302_2167395.shtml | Mar. 2,<br>2020 | Apr. 14,<br>2020 |
| Mar. 2,<br>2020 | 232863 | http://wjw.hubei.gov.cn/bmdt/ztzl/fkxxgzbdgrfyyq/xxfb/202003/t20200303_2171106.shtml | Mar. 3,<br>2020 | Apr. 14,<br>2020 |
| Mar. 3,<br>2020 | 179315 | http://wjw.hubei.gov.cn/bmdt/ztzl/fkxxgzbdgrfyyq/xxfb/202003/t20200304_2171824.shtml | Mar. 4,<br>2020 | Apr. 14,<br>2020 |
| Mar. 4,<br>2020 | 169180 | http://wjw.hubei.gov.cn/bmdt/ztzl/fkxxgzbdgrfyyq/xxfb/202003/t20200305_2172547.shtml | Mar. 5,<br>2020 | Apr. 14,<br>2020 |
| Mar. 5,<br>2020 | 162180 | http://wjw.hubei.gov.cn/bmdt/ztzl/fkxxgzbdgrfyyq/xxfb/202003/t20200306_2173603.shtml | Mar. 6,<br>2020 | Apr. 14,<br>2020 |
| Mar. 6,<br>2020 | 157115 | http://wjw.hubei.gov.cn/bmdt/ztzl/fkxxgzbdgrfyyq/xxfb/202003/t20200307_2174367.shtml | Mar. 7,<br>2020 | Apr. 14,<br>2020 |
| Mar. 7,<br>2020 | 148721 | http://wjw.hubei.gov.cn/bmdt/ztzl/fkxxgzbdgrfyyq/xxfb/202003/t20200308_2175120.shtml | Mar. 8,<br>2020 | Apr. 14,<br>2020 |
| Mar. 8,<br>2020 | 164137 | http://wjw.hubei.gov.cn/bmdt/ztzl/fkxxgzbdgrfyyq/xxfb/202003/t20200309_2175797.shtml | Mar. 9,<br>2020 | Apr. 14,<br>2020 |
| Mar. 9,<br>2020 | 142180 | http://wjw.hubei.gov.cn/bmdt/ztzl/fkxxgzbdgrfyyq/xxfb/202003/t20200310_2176880.shtml | Mar. 10,<br>2020 | Apr. 14,<br>2020 |
| Mar. 10,<br>2020 | 142100 | http://wjw.hubei.gov.cn/bmdt/ztzl/fkxxgzbdgrfyyq/xxfb/202003/t20200311_2178362.shtml | Mar. 11,<br>2020 | Apr. 14,<br>2020 |
| Mar. 11,<br>2020 | 66030 | http://wjw.hubei.gov.cn/fbjd/tzgg/202003/t20200312_2179602.shtml | Mar. 12,<br>2020 | Apr. 14,<br>2020 |
| Mar. 12,<br>2020 | 76510 | http://wjw.hubei.gov.cn/bmdt/ztzl/fkxxgzbdgrfyyq/xxfb/202003/t20200313_2180464.shtml | Mar. 13,<br>2020 | Apr. 14,<br>2020 |
| Mar. 13,<br>2020 | 61500 | http://wjw.hubei.gov.cn/bmdt/ztzl/fkxxgzbdgrfyyq/xxfb/202003/t20200314_2181224.shtml | Mar. 14,<br>2020 | Apr. 14,<br>2020 |
| Mar. 14,<br>2020 | 63593 | http://wjw.hubei.gov.cn/bmdt/ztzl/fkxxgzbdgrfyyq/xxfb/202003/t20200315_2181850.shtml | Mar. 15,<br>2020 | Apr. 14,<br>2020 |
| Mar. 15,<br>2020 | 54150 | http://wjw.hubei.gov.cn/bmdt/ztzl/fkxxgzbdgrfyyq/xxfb/202003/t20200316_2182355.shtml | Mar. 16,<br>2020 | Apr. 14,<br>2020 |
| Mar. 16,<br>2020 | 49785 | http://wjw.hubei.gov.cn/bmdt/ztzl/fkxxgzbdgrfyyq/xxfb/202003/t20200317_2183112.shtml | Mar. 17,<br>2020 | Apr. 14,<br>2020 |
| Mar. 17,<br>2020 | 57420 | http://wjw.hubei.gov.cn/bmdt/ztzl/fkxxgzbdgrfyyq/xxfb/202003/t20200318_2184169.shtml | Mar. 18,<br>2020 | Apr. 14,<br>2020 |

## References

1. Li Q, Guan X, Wu P, et al. Early Transmission Dynamics in Wuhan, China, of Novel Coronavirus-Infected Pneumonia. N Engl J Med 2020; 382(13): 1199-207.

2. Huang C, Wang Y, Li X, et al. Clinical features of patients infected with 2019 novel coronavirus in Wuhan, China. Lancet 2020; 395(10223): 497-506.

3. Chen N, Zhou M, Dong X, et al. Epidemiological and clinical characteristics of 99 cases of 2019 novel coronavirus pneumonia in Wuhan, China: a descriptive study. Lancet 2020; 395(10223): 507-13.

4. Zhu N, Zhang D, Wang W, et al. A Novel Coronavirus from Patients with Pneumonia in China, 2019. N Engl J Med 2020; 382(8): 727-33.

5. Wu F, Zhao S, Yu B, et al. A new coronavirus associated with human respiratory disease in China. Nature 2020; 579(7798): 265-9.

6. Lu R, Zhao X, Li J, et al. Genomic characterisation and epidemiology of 2019 novel coronavirus: implications for virus origins and receptor binding. Lancet 2020; 395(10224): 565-74.

7. Coronaviridae Study Group of the International Committee on Taxonomy of V. The species Severe acute respiratory syndrome-related coronavirus: classifying 2019-nCoV and naming it SARS-CoV-2. Nat Microbiol 2020; 5(4): 536-44.

8. World Health Organization. Coronavirus disease (COVID-19) Situation Dashboard. https://covid19.who.int/ (accessed April 18 2020).

9. Omer SB, Malani P, Del Rio C. The COVID-19 Pandemic in the US: A Clinical Update. JAMA 2020.

10. Phelan AL, Katz R, Gostin LO. The Novel Coronavirus Originating in Wuhan, China: Challenges for Global Health Governance. JAMA 2020.

11. Pan A, Liu L, Wang C, et al. Association of Public Health Interventions With the Epidemiology of the COVID-19 Outbreak in Wuhan, China. JAMA 2020.

12. Liu W, Yue XG, Tchounwou PB. Response to the COVID-19 Epidemic: The Chinese Experience and Implications for Other Countries. Int J Environ Res Public Health 2020; 17(7).

13. Adams JG, Walls RM. Supporting the Health Care Workforce During the COVID-19 Global Epidemic. JAMA 2020.

14. Hu Z, Song C, Xu C, et al. Clinical characteristics of 24 asymptomatic infections with COVID-19 screened among close contacts in Nanjing, China. Sci China Life Sci 2020.

15. Tanne JH, Hayasaki E, Zastrow M, Pulla P, Smith P, Rada AG. Covid-19: how doctors and healthcare systems are tackling coronavirus worldwide. BMJ 2020; 368: 1090.

16. Wu Z, McGoogan JM. Characteristics of and Important Lessons From the Coronavirus Disease 2019 (COVID-19) Outbreak in China: Summary of a Report of 72314 Cases From the Chinese Center for Disease Control and Prevention. JAMA 2020.

17. Novel Coronavirus Pneumonia Emergency Response Epidemiology Team. The epidemiological characteristics of an outbreak of 2019 novel coronavirus diseases (COVID-19) in China. Zhonghua Liu Xing Bing Xue Za Zhi 2020; 41(2): 145-51.

18. WHO Expert Committee on Biological Standardization. Guidelines on viral inactivation and removal procedures intended to assure the viral safety of human blood plasma products. 2004.

19. Xinhua News. Backgrounder: Nationwide aid to Hubei’s battle against coronavirus. 2020. http://www.xinhuanet.com/english/2020-03/17/c_138888353.htm (accessed April 14 2020).

20. Chan JF, Yuan S, Kok KH, et al. A familial cluster of pneumonia associated with the 2019 novel coronavirus indicating person-to-person transmission: a study of a family cluster. Lancet 2020; 395(10223): 514-23.

21. Leung NH, Chu DK, Shiu EY, et al. Respiratory virus shedding in exhaled breath and efficacy of face masks. Nat Med 2020.

22. Wang D, Hu B, Hu C, et al. Clinical Characteristics of 138 Hospitalized Patients With 2019 Novel Coronavirus-Infected Pneumonia in Wuhan, China. JAMA 2020.

23. Ong SWX, Tan YK, Chia PY, et al. Air, Surface Environmental, and Personal Protective Equipment Contamination by Severe Acute Respiratory Syndrome Coronavirus 2 (SARS-CoV-2) From a Symptomatic Patient. JAMA 2020.

24. Grasselli G, Zangrillo A, Zanella A, et al. Baseline Characteristics and Outcomes of 1591 Patients Infected With SARS-CoV-2 Admitted to ICUs of the Lombardy Region, Italy. JAMA 2020.

